# CRABP2 Upregulation in Perichondral Stem Cells is Associated with Microtia

**DOI:** 10.64898/2025.12.18.25342452

**Authors:** Jingheng Zhang, Xuzhong Hu, Xinghua Yang, Shaorong Lei, Leren He, Haiyue Jiang, Lin Lin, Dingyu Wu

**Affiliations:** Chinese Academy of Medical Sciences & Peking Union Medical College Plastic Surgery Hospital and Institute, Beijing, 100144, China; Department of Plastic Surgery, Xiangya Hospital, Central South University, Changsha, 410008, China

**Keywords:** Microtia, Etiology, Elastic cartilage, CRABP2

## Abstract

Microtia is a common congenital craniofacial malformation characterized by the partial or complete absence of the external ear structure. Despite its relatively high incidence, the pathogenesis of microtia remain poorly understood. In this study, we analyzed both single-cell and bulk RNA sequencing data from microtia cases and identified a population of COL1^+^HES1^+^ mesenchymal stem cell in perichondrium with significantly higher expression of the CRABP2 gene, a gene that encodes a nuclear transporter of retinoic acid. Gene expression analysis further confirmed that the RA signaling intensity and stemness are both higher in COL1^+^HES1^+^ perichondral stem cells from microtia patients, possibly due to elevated CRABP2 levels. Through histological verification we further confirmed the presence of this cell population with high CRABP2 expression in the perichondrium. Mechanistically, the elevated CRABP2 expression in perichondral stem cells seen in microtia patients may cause dysregulated RA signaling and disrupt the regulation of stem cell differentiation during auricular development. Histological analysis further revealed higher KLF2 expression as well as cartilage hypoplasia in microtia samples. Our study identified that the CRABP2-induced RA dysregulation in COL1^+^HES1^+^ perichondral stem cells may contribute to microtia. These findings offer new insights into the etiology of microtia and provide potential directions for prenatal prevention and tissue engineering treatments.

## Introduction

Microtia is a common congenital craniofacial malformation characterized by the partial or complete absence of the external ear structure^1^. The prevalence of microtia varies significantly across the globe, with higher incidence rates observed in regions such as East Asia and Latin America compared to Western populations^2,3^. The severity of microtia is typically classified into four grades, ranging from slight reduction in ear size (Grade I) to complete absence of the ear (Grade IV, also known as anotia), with Grade III being the most common. Given the prominent position of the external ear on the face, microtia not only affects hearing but also poses significant psychosocial challenges for patients, such as anxiety, depression, and diminished self-esteem^4^. Moreover, the condition is frequently associated with external auditory canal atresia, leading to varying degrees of conductive hearing loss, which in turn can delay speech and language development^5^.

Despite its relatively high incidence, the etiology and pathogenesis of microtia remain poorly understood. It is currently believed that microtia may be related to abnormal differentiation of chondrogenic stem cells during embryonic development^6–8^. In the early stages of embryonic development, the auricle forms from mesenchymal tissue derived from the first branchial arch. Mesenchymal stem cells differentiate into chondral progenitor cells, which then construct the cartilage framework of the auricle. However, the mechanisms underlying abnormal cartilage development seen in microtia is still unclear^9^.

Previous study has identified retinoic acid (RA) as a crucial regulator in cartilage development^10,11^. Strong expression of RAR-beta, a nuclear receptor of RA, has been found in the perichondrium of chick embryos^12^. Also, studies on chicken sternal chondrocytes have reported that within 4 days of RA treatment, chondrocytes stop synthesizing the cartilage-specific type II collagen and turn on the synthesis of types I and III collagen and fibronectin^13^. In humans, accidental ingestion of RA during pregnancy can cause spontaneous abortions, major congenital anomalies^14^ and auricular malformations^15,16^. Therefore, the potential role of RA signaling in microtia has been a focal point for researchers^17^, due to the observed resemblance between isotretinoin embryopathy and microtia^18^. Whole-exome sequencing (WES) has revealed nonsense mutations in CYP26A1 and CYP26B1, two catabolize enzymes of RA, in microtia patients^19,20^.

In this study, we analyzed both single-cell^21^ and bulk sequencing data^22^ from microtia cases and identified a subpopulation of mesenchymal stem cells with significantly higher expression of the CRABP2 gene. CRABP2 encodes a key transporter involved in RA metabolism. Our research further confirmed through histological verification the presence of a population of COL1^+^HES1^+^ stem cells with high CRABP2 expression in the perichondrium. This finding not only provides new insights into the molecular mechanisms underlying microtia but also offers potential targets for future preventive and therapeutic strategies.

## Methods

### Data acquisition and processing

For both RNA-seq (GSE227119) and scRNA-seq (GSE179135) data, the accession number of corresponding samples was retrieved from the Sequence Read Archive (SRA) database, and data was downloaded with the sratoolkit (version 3.1.1). The analysis was performed on a rack-mount server with a Linux operating system (Ubuntu 22.04). The RNA-seq dataset comprised of 3 normal samples, 3 samples from 2^nd^ degree NSM and 3 samples from 3^rd^ degree NSM patients. The scRNA-seq dataset comprised of 6 normal samples and 3 samples from 3^rd^ degree NSM patients.

### RNA-seq data analysis

The RNA-seq data was trimmed with trim_galore (version 0.6.7), mapped to reference genome Hg38 with STAR (version 2.7.10b) and quantified with featurecounts (version 2.0.5) to obtain the raw count matrix. Then, differential expression analysis was performed with DESeq2 (version 1.38.3) to obtain a list of differentially expressed genes. WGCNA was conducted with the cloud-based platform https://bic.ac.cn to obtain gene modules.

### scRNA-seq data analysis

The scRNA-seq data was quantified with Cellranger (version 7.0.1) to obtain h5ad and bam files. The h5ad files served as of input of Scanpy (version 1.9.3) to conduct clustering. The bam files were used to perform trajectory inference by utilizing velocyto (version 0.17.17) combined with scvelo (version 0.2.5).

### Sample collection and formalin-fixed paraffin-embedded (FFPE) tissue sample preparation

This study complied with the Declaration of Helsinki and was approved by the ethics committee of Xiangya Hospital, Central South University (#2024121653). Written informed consent was obtained from all participants. Sample cartilage was obtained from four 3^rd^ degree microtia patients underwent auricular reconstruction surgery and four healthy individuals underwent rhinoplasty procedures. For microtia patients, the degree of auricular deformity was determined under Marx classification by an experienced plastic surgeon. Patients included were all non-syndromic microtia (NSM) patients absent of additional abnormalities.

Once harvested, samples were rinsed with 0.9% normal saline, immersed into 4% paraformaldehyde (PFA) and fixed overnight. Then, samples were dehydrated, paraffinized and sectioned.

### RNA extraction and quantitative PCR of FFPE samples

RNA extraction was performed with miRNeasy FFPE kit (217504, Qiagen, Germany) according to manufacturer’s instructions. Following RNA elution, the concentration of RNA was measured with a Nanodrop One microvolume spectrophotometer. Then, reverse transcription of RNA into cDNA was performed with PrimeScript RT reagent kit (RR036A, Takara, Japan), and amplified with SYBR Green 1 master (4707516001, Roche, Switzerland) on a Lightcycler 96 real-time PCR instrument. Three-step amplification protocols were used.

### Immunofluorescence staining of FFPE samples

Firstly, antigen retrieval of FFPE sections were performed at 98 LJ for 10 mins with Ph-9.0 Tris-EDTA antigen retrieval buffer (G1203, Servicebio, China). Then, Sections were blocked with 5% BSA/PBS for 1 hour and incubated with COL-1 and HES1 antibodies in a cold room overnight. Then, sections were extensively washed with PBS, and incubated with secondary antibodies at room temperature for 1 hour. Then, after extensively washed, sections were incubated with CoraLite 594-conjugated CRABP2 at room temperature for 1 hour. Finally, sections were washed, counter-stained with DAPI, and imaged. The following primary antibodies and dilutions were used: COL-1 (1:150, 67288-1-Ig, Proteintech, China), CoraLite 594-conjugated CRABP2 (1:75, CL594-10225, Proteintech, China), and HES1 (1:200, 11988, CST, United States). The following secondary antibodies and dilutions were used: CoraLite Plus 488-Goat Anti-Mouse Recombinant Secondary Antibody (1:300, RGAM002, Proteintech, China) and CoraLite Plus 647-Goat Anti-Rabbit Recombinant Secondary Antibody (1:300, RGAR005, Proteintech, China). Then, sections were extensively washed, mounted and imaged with a laser scanning confocal microscope (AXR, Nikon, Japan).

### Immunohistochemistry (IHC) of FFPE tissue samples

Tissue sections were subject to deparaffinization and rehydration. Then, sections were treated with Tris-EDTA antigen retrieval buffer (pH 9.0) at 98 LJ for 10 mins, blocked with 5% BSA/PBS at room temperature for 1 hour, and treated with 3% H_2_O_2_ for 15 mins. Then, sections were incubated with primary antibody at room temperature for 1 hour, extensively washed, and incubated with HRP-conjugated secondary antibody at room temperature for 15 mins. After washing, DAB substrate was added to sections, and sections were counterstained with hematoxylin. After washing and dehydration, sections were mounted and air dried. KLF2 primary antibody (A16480, Abclonal, China) and HRP-conjugated Goat Anti-Rabbit secondary antibody (SA00001-2, Proteintech, China) were used. The IHC slides was scanned with a panoramic midi scanner (3DHistech) and staining intensities was quantified with the ImageJ plugin IHC Profiler as channel deconvoluted images.

## Results

### CRABP2 mRNA levels correlate positively with microtia severity

Since the severity of auricular hypoplasia is markedly different among microtia patients, we first aimed to identify genes whose expression patterns are positively correlated with the severity of microtia. We utilized dataset GSE227119, which comprises RNA-seq data from auricular cartilage of patients with mild and severe microtia, as well as normal controls (Supplementary table 1). Firstly, we performed differential expression analysis among groups (mild vs. normal and severe vs. normal), and extracted 113 overlapping genes that were upregulated in both mild and severe microtia (Figure 1A). To prioritize these genes, we calculated the difference in log2FoldChange values between the severe and mild groups (log2FC (severe vs. normal) minus log2FC (mild vs. normal), denoted as Delta(log2FC)). Using this approach, we identified top 10 genes with higher expression in the severe group compared to the mild group (Figure 1B). Secondly, as an alternative method, we conducted weighted correlation network analysis (WGCNA) on this dataset to identify gene modules that are positively correlated to microtia severity. The WGCNA identified 13 gene modules, of which only one exhibited a statistically significant positive correlation with disease severity (Figures 1C and 1D). This module contained 272 genes. By intersecting the 113 genes from differential expression analysis and the 272 genes from WGCNA, we obtained a refined set of 26 candidate genes (Figure 1E), including 16 protein-coding genes, 3 long non-coding RNAs, and 7 other non-coding features. To gain functional insights, we performed Gene Ontology (GO) enrichment analysis, which revealed significant enrichment in pathways related to embryonic morphogenesis and retinoic acid signaling (Figure 1F). To validate these findings, we collected auricular cartilage samples from microtia patients as well as healthy individuals, and performed quantitative PCR against the 16 coding genes in this 26-gene set (Figure 1G). Eight genes, namely CRABP2, RGS4, ALDH1A3, AQP7, OSR1, CDCP1, SYT7, and TMEM119 showed up-regulation consistent with our bioinformatic analysis.

**Figure 1.**
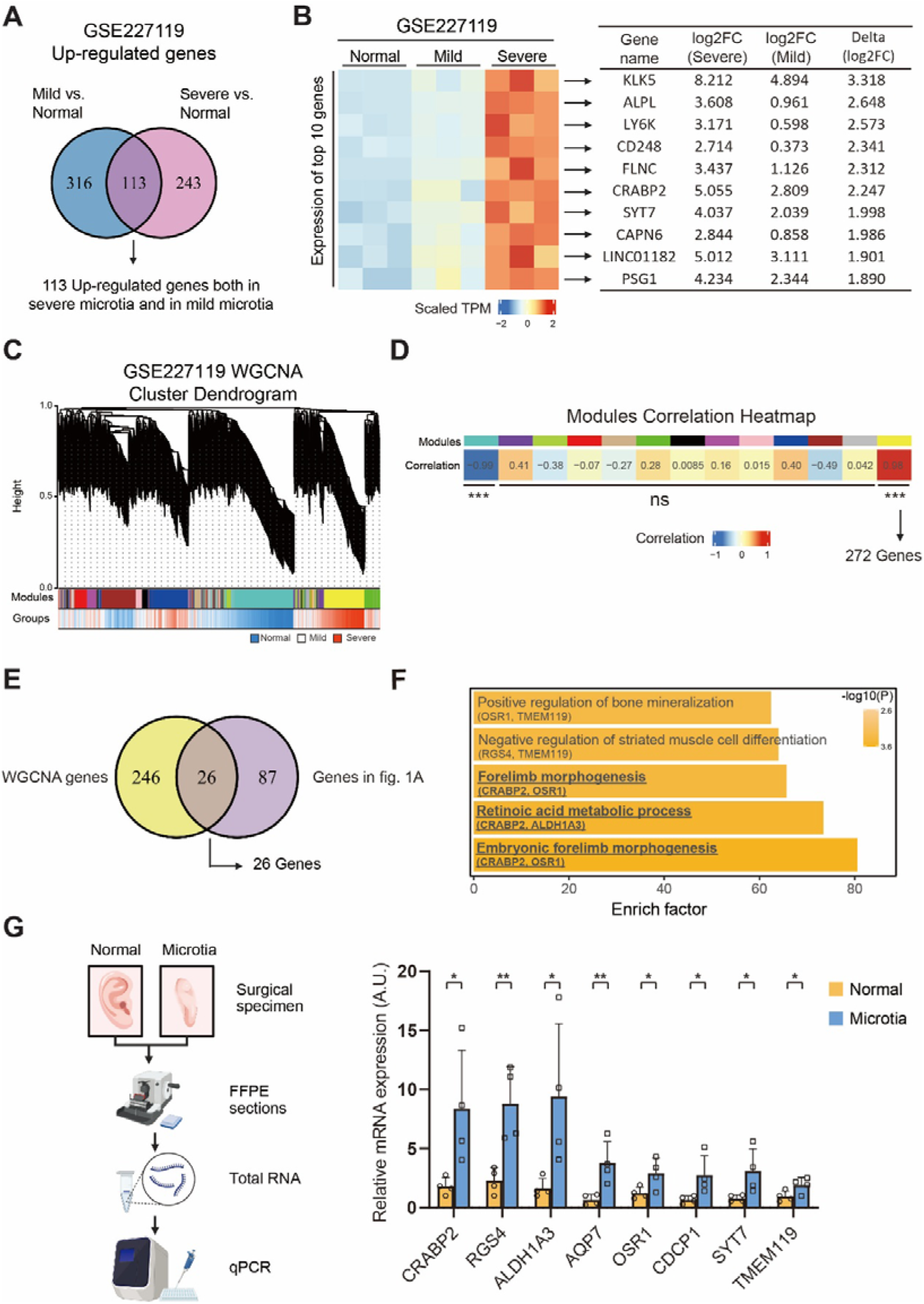
Identification of 26 genes that positively correlate with microtia severity. (A) Venn diagram showing the 113 overlapping genes of two sets of differential expression analysis. (B) Heatmap and table of top 10 genes among the 113 overlapping genes showing in (A). (C) Waterfall plot demonstrating the 13 gene modules identified by WGCNA. (D) Correlation and p values of each module shown in (C). (E) Venn diagram showing the 26 overlapping genes that occurred both in differential expression analysis and in WGCNA. (F) Gene ontology enrichment results of the 26 genes showed in (E). (G) Quantitative PCR validation of the 16 protein-coding genes in the 26-gene list. Only genes with significant up-regulation in qPCR results was showed. *P<0.05, **P<0.01, Student’s t test.

### CRABP2 is specifically expressed in COL1^+^HES1^+^ perichondral stem cells

Next, we aimed to elucidate the relationship between CRABP2 up-regulation and the pathogenesis of microtia. To this end, we analyzed dataset GSE179135^6^, a set of scRNA-seq data comprising auricular cartilage samples from microtia patients and healthy individuals. After UMAP dimension reduction and Leiden clustering, 12 cell clusters were obtained with their respective marker genes (Figures 2A and 2B). Cell identities were assigned based on the expression of established lineage- and cell type-specific markers. Based on expression of COL2A1 and ACAN, clusters 0, 1, 2, 3, 5 and 7 were collectively assigned to the chondrocyte lineage (Figures 3C). Based on expression of COL1A1 and VCAN, clusters 4 and 9 were assigned to the perichondral cell lineage (Figures 3C). Cluster 6 was assigned to pericytes based on the expression of ACTA2. Cluster 8 was assigned to endothelial cells based on the expression of CLDN5. Cluster 10 was assigned to immune cells based on the expression of PTPRC/CD45. Among these cell clusters, cluster 9 caught our attention due to its position on UMAP projection lies between the chondrocyte lineage and the perichondral cell lineage. Examination of marker genes expressed by cluster 9 revealed high expression of multiple markers of human mesenchymal stem cells (hMSC), suggesting its identity as a chondroprogenitor population.

**Figure 2.**
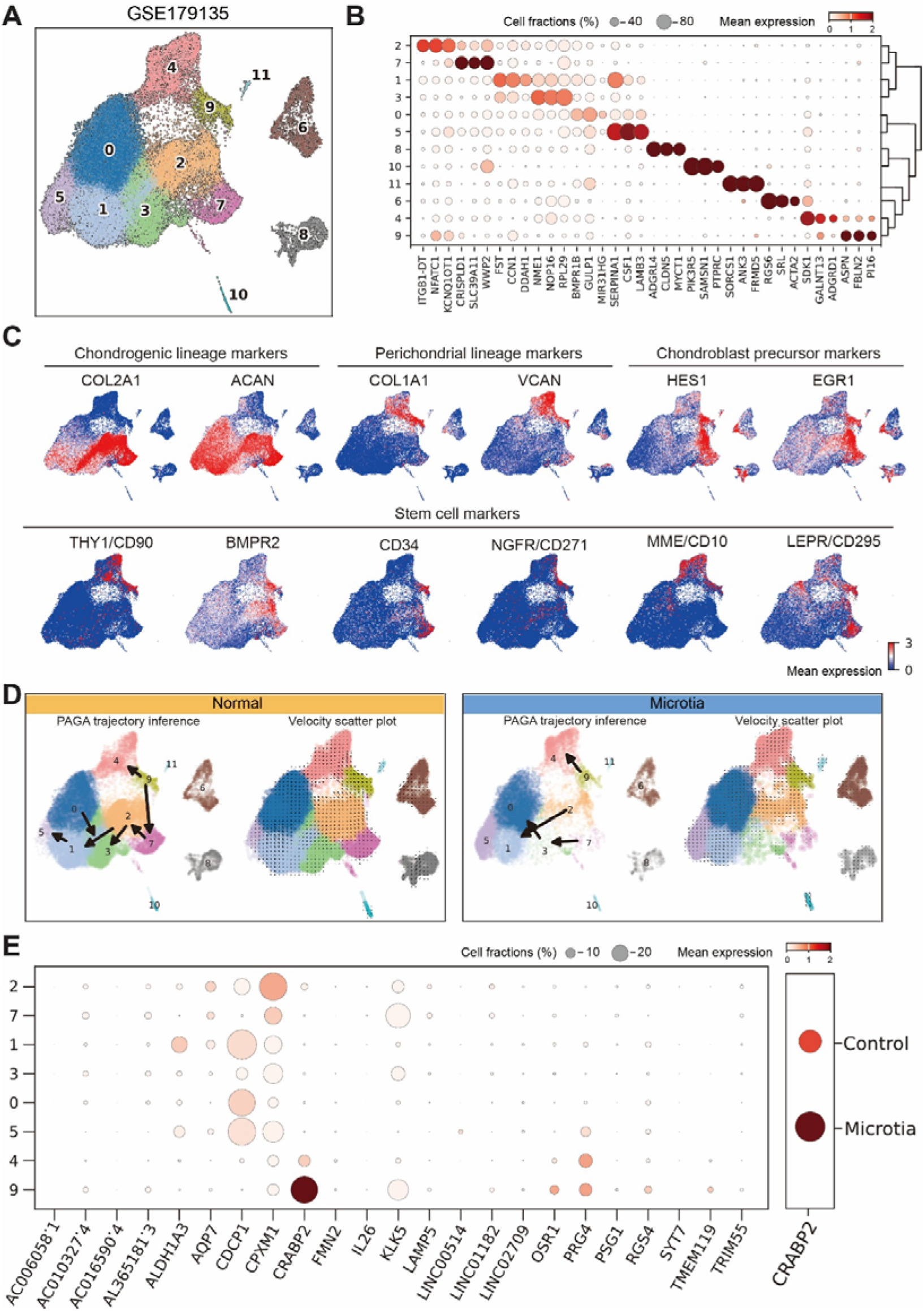
Specific expression of CRABP2 in COL1^+^HES1^+^ perichondral stem cells. (A) UMAP projection and unbiased clustering of all cells in GSE179135 derived 12 cell clusters. (B) Dot plot shows the selected marker genes of each cell cluster. (C) Expression of selected lineage-specific genes in all cells highlighting three lineages. (D) Trajectory inference and velocity plot of all cells, split by sample type. (E) Dot plot shows the expression pattern of 26 genes in all cell clusters.

**Figure 3.**
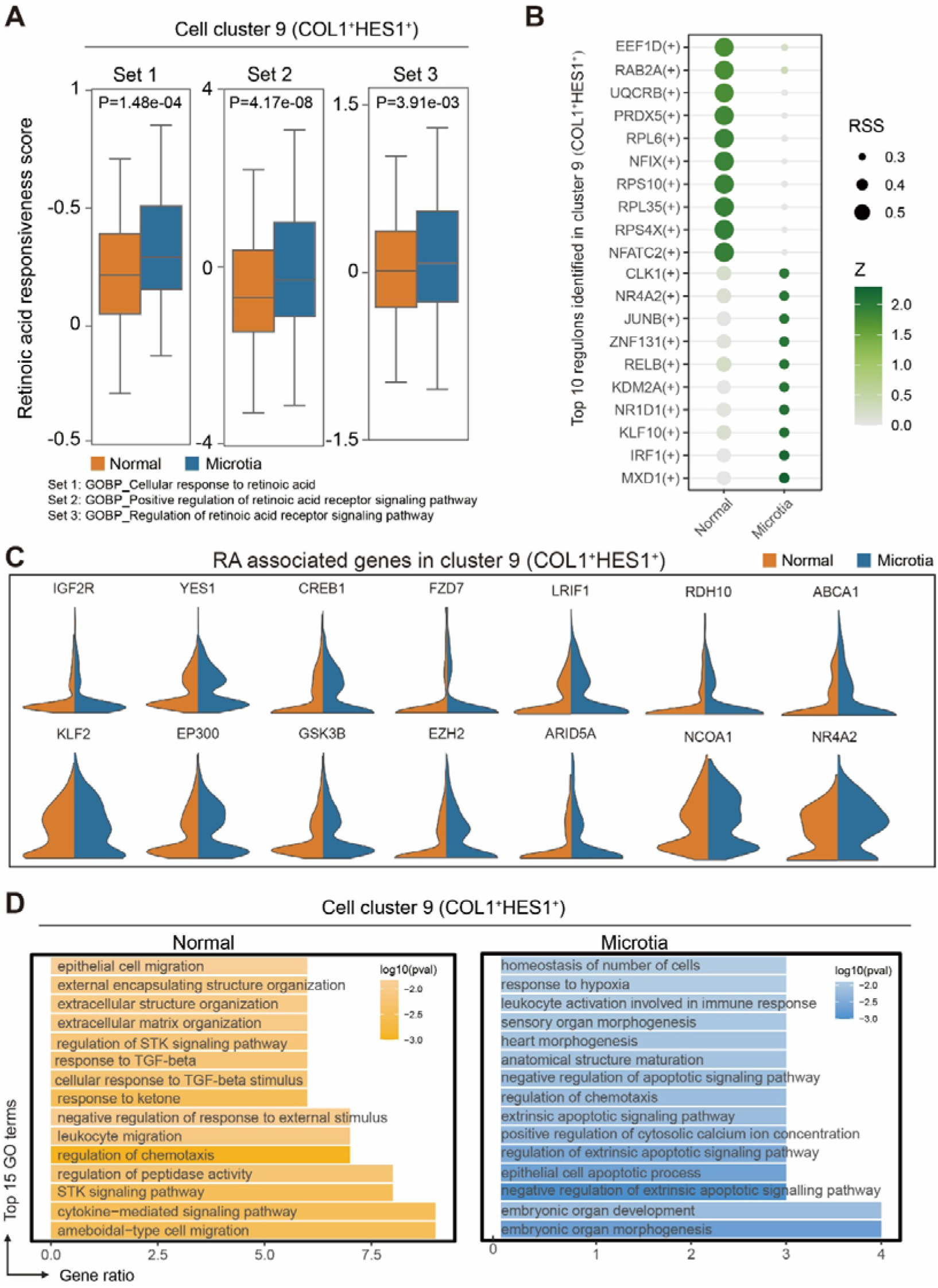
Elevated Retinoic acid signaling intensity and stemness in COL1+HES1+ perichondral stem cells from microtia patients. (A) Boxplot shows the RA responsiveness score measured by three different gene sets. (B) Dot plot shows the top 10 active regulons predicted by pyscenic in the COL1^+^HES1^+^ perichondral stem cells from normal and microtia samples respectively. (C) Gene ontology enrichment results of genes up-regulated in COL1^+^HES1^+^ perichondral stem cells from normal samples (left) and in microtia samples (right). (D) Expression level of RA associated genes in COL1^+^HES1^+^ perichondral stem cells from normal samples (left) and in microtia samples (right). ***P<0.001, Student’s t test.

In normal samples, trajectory inference based on RNA velocity revealed two differentiation routes (Figure 2D). Both routes originated from cluster 9: one route representing perichondral lineage (cluster 9 to cluster 4) and the other representing the chondrogenic lineage (cluster 9 to clusters 7, 2, 3, and 1, culminating in cluster 5). However, in microtia samples, while the perichondral lineage remained intact, the chondrogenic lineage exhibited dysregulation, potentially explained the insufficient auricular development observed in microtia patients.

To further investigate the cellular basis of the 26 genes, we examined their expression at single cell resolution, which showed a distinct cell type-specific expression pattern of CRABP2 in cluster 9 (Figure 2E).

### RA signaling intensity and stemness are both higher in COL1^+^HES1^+^ perichondral stem cells from microtia patients

Given that CRABP2 functions as a transporter of retinoic acid (RA), facilitating its translocation from the cytoplasm to the nucleus, we hypothesized that RA signaling activity might be elevated in microtia. Indeed, by quantifying RA signal in cells using three different gene sets from Gene Ontology (GO), the results confirmed that the signal intensity of RA pathway is elevated in cluster 9 cells from microtia patients compared to normal individuals (Figure 3A). On the gene level, multiple genes associated with RA signaling were upregulated in cluster 9 cells derived from microtia patients (Figure 3C).

RA signaling is one of the key regulators of cell stemness. Thus, we hypothesized that heightened RA signaling in cluster 9 may promote the maintenance of cell stemness. Regulon analysis further revealed that the transcription factors in cluster 9 cells from microtia exhibited stronger cell stemness (Figure 3B). Specifically, MXD1 and IRF1 maintains cell stemness, and KLF10 inhibit hMSC chondrogenesis. In contrast, transcription factors in cluster 9 cells from normal samples were associated with chondrogenic lineage commitment. Notably, NFATC2 regulates cartilage homeostasis, NFIX controls chondrocyte extracellular matrix (ECM) synthesis, and PRDX5 has been linked to chondrocyte growth. Consistent with these findings, GO analysis on differential expressed genes (microtia group vs. normal group of the cluster 9) revealed that genes associated with cell migration and ECM organization were upregulated in normal samples, whereas genes related to embryonic organ morphogenesis were upregulated in microtia samples, suggesting that cluster 9 cells in microtia remain in a more primitive developmental state (Figure 3D).

### Proportion of CRABP2^+^ cells in COL1^+^HES1^+^ perichondral stem cells is higher in microtia patients

To spatially resolve the quantity and distribution of CRABP2^+^ cells within COL1^+^HES1^+^ perichondral stem cells in auricular cartilage, we performed immunofluorescence staining on auricular tissue sections from microtia patients and normal individuals. The results validated that CRABP2^+^COL1^+^HES1^+^ cells were predominantly localized within the perichondrium of microtia samples (Figures 4A, 4B and 4C).

**Figure 4.**
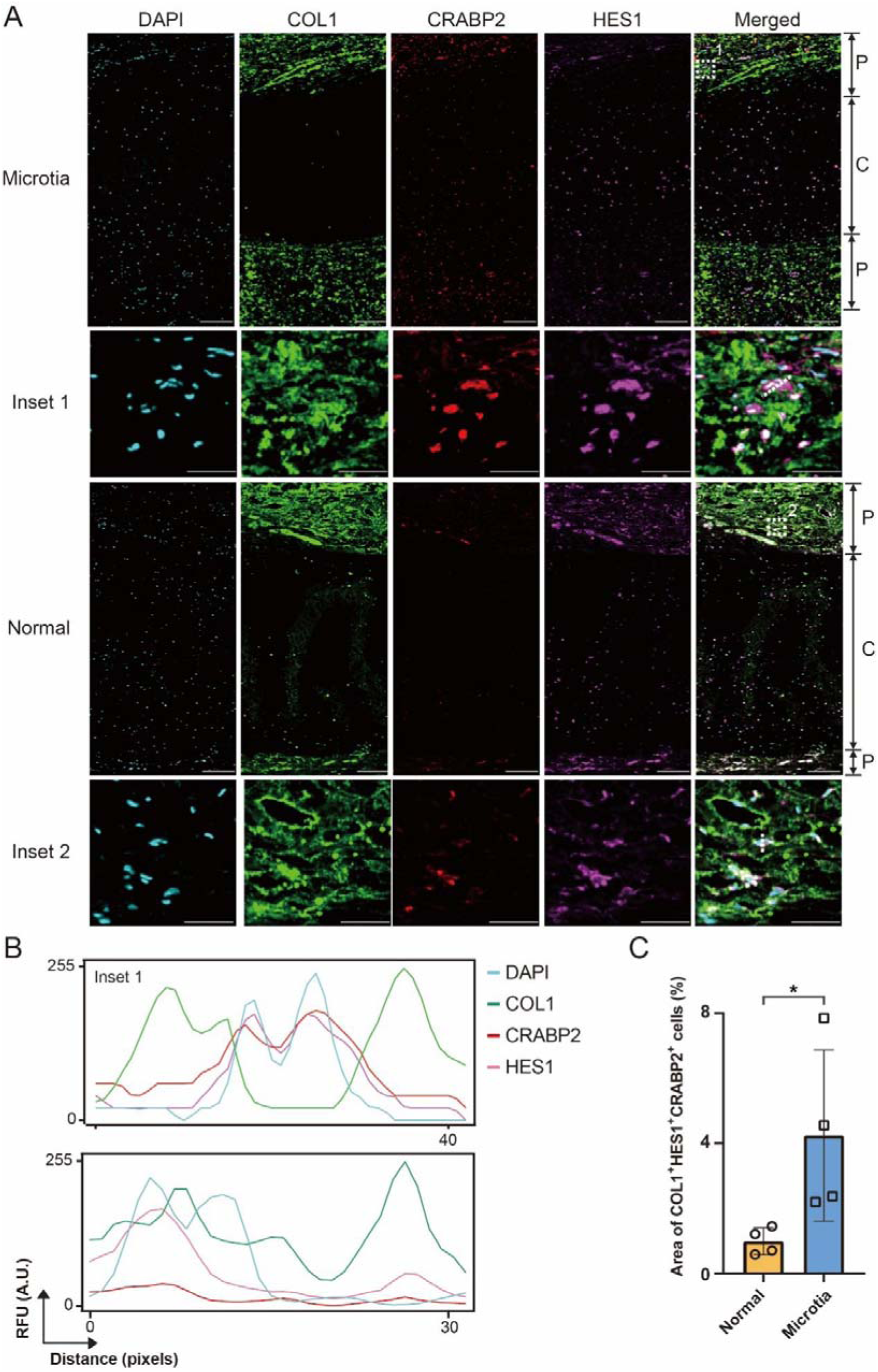
Enrichment of COL1^+^HES1^+^ perichondral stem cells in the perichondrium of microtia samples. (A) Representative immunofluorescence staining images of auricular cartilage slices from microtia and normal samples. Scale bar for large images: 150 µm. Scale bar for insets: 50 µm. (B) Line intensity profiles of DAPI, anti-COL1, anti-CRABP2 and anti-HES1 immunostainings. (C) Boxplot shows the quantification result of CRABP2^+^COL1^+^HES1^+^ proportions among COL1^+^HES1^+^ perichondral stem cells in normal and microtia samples. *P<0.05, Student’s t test.

### Cartilage hypoplasia is observed in microtia patients

Given the observed elevation of RA signaling intensity and cell stemness was observed in COL1^+^HES1^+^ perichondral stem cells within auricular cartilage, we sought to validate these findings through histology. We performed immunohistochemistry staining of KLF2, a RA target gene that inhibit stem cell differentiation^23^, and discovered that KLF2 is over-expressed in the auricular cartilage from microtia patients compared to that of normal individuals (Figure 5A).

**Figure 5.**
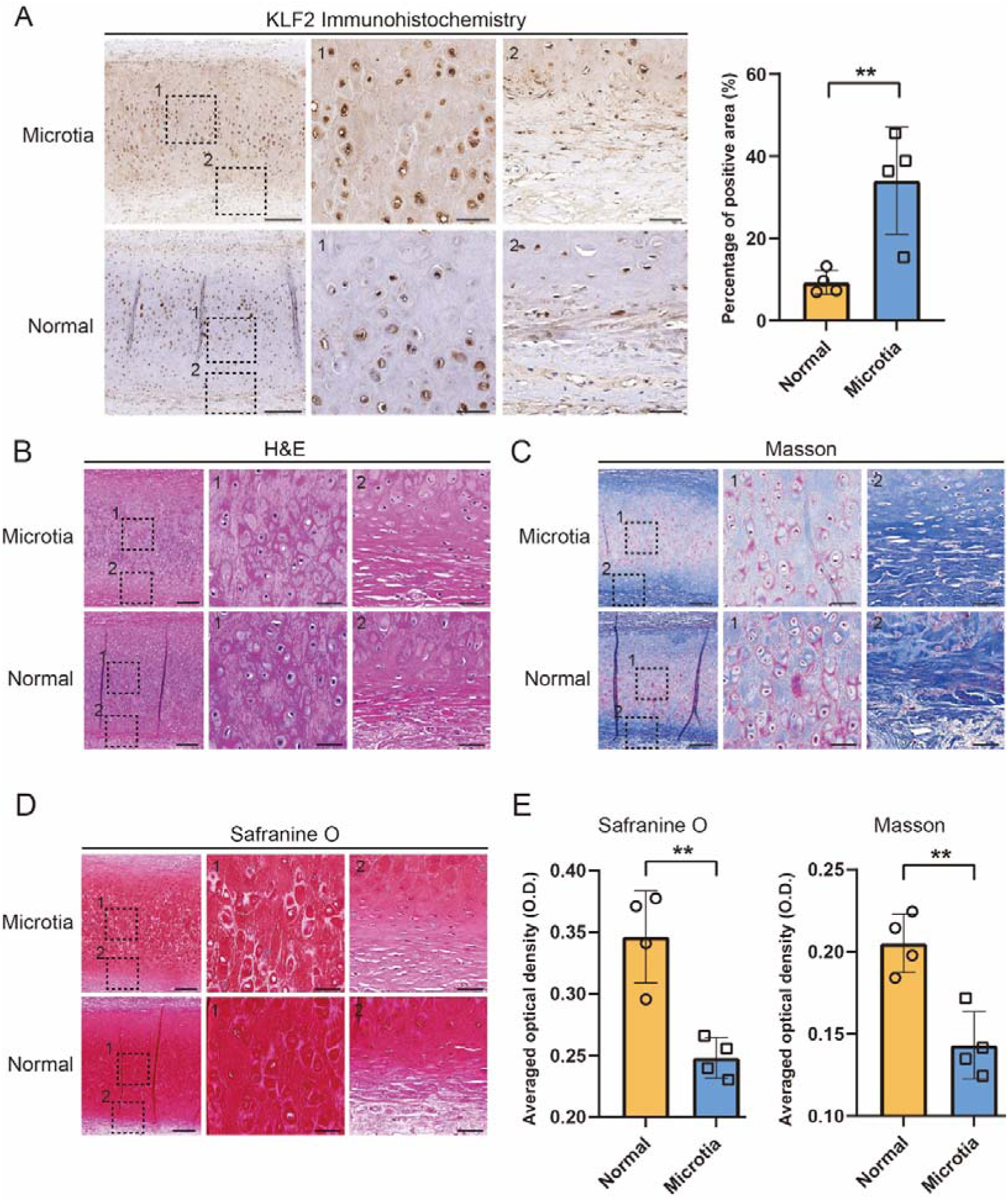
Cartilage hypoplasia is observed in microtia patients. (A) Representative immunohistochemistry images(left) and quantification results (right) of KLF2. Scale bar for large images: 500 µm. Scale bar for insets: 20 µm. (B) Representative H&E staining, (C) Masson trichrome staining, and (D) Safranine O staining of auricular cartilage slices from normal and microtia samples. Scale bar for large images: 200 µm. Scale bar for insets: 50 µm. (E) Quantification results of Safranine O (left) and Masson trichrome staining (right). **P<0.01, Student’s t test.

**Figure 6.**
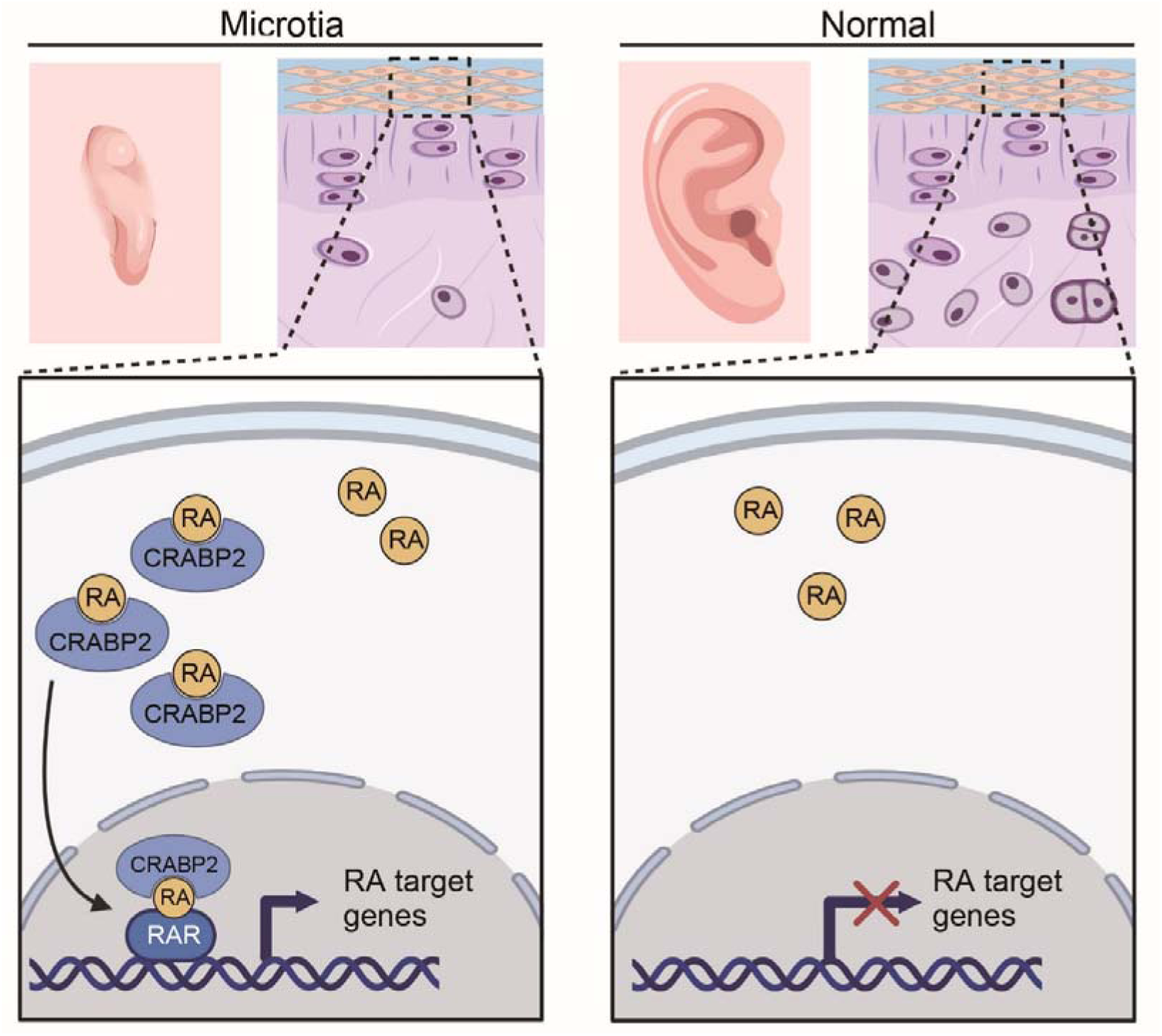
Schematic Diagram of the working model of this study.

Additionally, we conducted histological analysis staining to assess cartilage composition. Masson trichrome staining revealed that the staining of Ponceau S, which linked to protean content in microtia sections, exhibited significantly weaker than in normal sections (Figure 5C). We also performed the staining of Safranine O which is linked to the content of proteoglycan. The result also showed that the intensity of staining in microtia sections is significantly weaker than in normal sections (Figure 5D). This result revealed the hypoplasia of microtia cartilage, concurring with the elevated stemness in the auricular cartilage from microtia patients.

## Data Availability

All data produced in the present study are available upon reasonable request to the authors.

## Conflicts of Interest

The authors have no financial relationships or conflicts of interest to declare.

## Research funding

This work was supported by the National Key R&D Program of China (2024YFA1107800) and the Chinese Academy of Medical Sciences Innovation Fund for Medical Sciences (2021-I2M-1–052).

## Author Contributions

H.Y.J., L.L. and D.Y.W. designed and oversaw the study. J.H.Z., X.Z.H., D.Y.W. and L.R.H. performed the experiments and wrote the manuscript. S.R.L. and X.H.Y. collected the clinical sample. All authors read and approved the manuscript.

---

**Supplementary Table 1.** Demographics data of patients included in this study. Age of patients were written in non-overlapping age ranges.

| Patient No. | Source | Accession number | Data | Age | Sex | Group |
| --- | --- | --- | --- | --- | --- | --- |
| 1 | GSE227119 | GSM7091921 | RNA-seq | - | - | Normal |
| 2 |  | GSM7091922 |  | - | - | Normal |
| 3 |  | GSM7091923 |  | - | - | Normal |
| 4 |  | GSM7091924 |  | - | - | MIC_II |
| 5 |  | GSM7091925 |  | - | - | MIC_II |
| 6 |  | GSM7091926 |  | - | - | MIC_II |
| 7 |  | GSM7091927 |  | - | - | MIC_III |
| 8 |  | GSM7091928 |  | - | - | MIC_III |
| 9 |  | GSM7091929 |  | - | - | MIC_III |
| 10 | GSE179135 | GSM5410484 | ScRNA-seq | 41-45 | F | Normal |
| 11 |  | GSM5410485 |  | 21-25 | M | Normal |
| 12 |  | GSM5410486 |  | 31-35 | F | Normal |
| 13 |  | GSM5410487 |  | 6-10 | F | Normal |
| 14 |  | GSM5410488 |  | 56-60 | F | Normal |
| 15 |  | GSM5410489 |  | 6-10 | F | Normal |
| 16 |  | GSM5410490 |  | 6-10 | M | MIC_III |
| 17 |  | GSM5410491 |  | 6-10 | M | MIC_III |
| 18 |  | GSM5410492 |  | 6-10 | M | MIC_III |
| 19 | This study | - | FFPE | 21-25 | F | Normal |
| 20 |  | - |  | 16-20 | F | Normal |
| 21 |  | - |  | 21-25 | M | Normal |
| 22 |  | - |  | 21-25 | M | Normal |
| 23 |  | - |  | 11-15 | F | MIC_III |
| 24 |  | - |  | 11-15 | F | MIC_III |
| 25 |  | - |  | 16-20 | M | MIC_III |
| 26 |  | - |  | 11-15 | M | MIC_III |

## Notes

### Competing Interest Statement

The authors have declared no competing interest.

### Author Declarations

This study complied with the Declaration of Helsinki and was approved by the ethics committee of Xiangya Hospital, Central South University (#2024121653). Written informed consent was obtained from all participants.

